# Development and validation of a novel *in-vivo* vascular injury score for prediction of in-stent restenosis

**DOI:** 10.1101/2023.03.22.23286988

**Authors:** Anne Cornelissen, Roberta Andreea Florescu, Stefanie Reese, Marek Behr, Anna Ranno, Kiran Manjunatha, Nicole Schaaps, Christian Böhm, Elisa Anamaria Liehn, Liguo Zhao, Pakhwan Nilcham, Andrea Milzi, Jörg Schröder, Felix Jan Vogt

**Author notes:** **Corresponding Author:** Dr. med. Anne Cornelissen University Hospital Aachen, Department of Cardiology, Angiology, and Internal Intensive Medicine Pauwelsstraße 30, 52074 Aachen, Germany. This author takes responsibility for all aspects of the reliability and freedom from bias of the data presented and their discussed interpretation. **Disclosures:** None.

## Abstract

**Background:** Despite optimizations of coronary stenting technology, a residual risk of in-stent restenosis (ISR) remains. Vessel wall injury has important impact on the development of ISR. While injury can be assessed in histology, there is no injury score available to be used in clinical practice.

**Methods:** Seven rats underwent abdominal aorta stent implantation. At 4 weeks after implantation, animals were euthanized, and strut indentation, defined as the impression of the strut into the vessel wall, as well as neointimal growth were assessed. Established histological injury scores were assessed to confirm associations between indentation and vessel wall injury. In addition, stent strut indentation was assessed by optical coherence tomography (OCT) in an exemplary clinical case.

**Results:** Stent strut indentation was associated with vessel wall injury in histology. Furthermore, indentation was positively correlated with neointimal thickness, both in the per-strut analysis (r=0.5579) and in the per-section analysis (r=0.8620; both p ≤0.001). In a clinical case, indentation quantification in OCT was feasible, enabling assessment of injury *in vivo*.

**Conclusion:** Assessing stent strut indentation enables periprocedural assessment of stent-induced damage *in vivo* and therefore allows for optimization of stent implantation. The assessment of stent strut indentation might become a valuable tool in clinical practice.

## Introduction

In-stent restenosis (ISR) is a gradual re-narrowing of a stented coronary artery which typically presents as recurrent angina and occurs mostly between 3 to 12 months after percutaneous coronary intervention (PCI) and stent placement. Although the introduction of drug-eluting stents (DES) has reduced the incidence of ISR, there is still an up to 10% risk of ISR following percutaneous coronary interventions (1, 2). The pathogenesis of ISR in DES is incompletely understood, however, multifarious risk factors have been associated with DES-ISR. While biological and genetic parameters often cannot be sufficiently modified with currently available medical approaches, procedural mechanical and technical factors have been shown to contribute to DES-ISR. Stent underexpansion has been associated with an increased risk of ISR and stent thrombosis (3, 4). Thus, for optimal results following stent implantation, a minimal stent area (MSA) of less than 80% of the average reference lumen area for non-left main lesions and of less than 90% of the reference lumen area for left main lesions should be achieved (5).

On the other hand, there is also a risk of damaging the vessel wall due to increased balloon pressure, and a correlation between stent-induced vessel injury and ISR has been described (6). Of note, damage to the arterial wall often is only apparent in histology whereas it is difficult to quantify clinically (7, 8). Hence, various scores for the histopathological assessment of stent-induced injury have been suggested, including the Schwartz score (9) and a score introduced by Waale (10). The Schwartz score assesses the presence or absence of ruptures of the elastic membranes whereas the Waale score assesses the angular deformation of the media by the stent struts, and the degree of media compression correlated with increased neointimal growth (10).

However, these scores have been developed for the *postmortem* evaluation of implanted stents as they assess features of vascular injury in histologically processed tissue. Thus, none of these scores is suitable for intraprocedural vascular injury assessment and scoring *in vivo*.

Ideally, one would aim for novel treatment strategies, individually tailored to the patient’s coronary anatomy. Identifying certain mechanical risk factors that are predictive for the occurrence of ISR could not only provide some insight into the process of ISR but could also lead to new treatment strategies for the prevention of ISR.

Optical coherence tomography (OCT) is a well-established high-resolution intravascular imaging modality to visualize coronary arteries with and without stent struts. Of note, OCT also allows for visualization and assessment of stent-induced injuries. Although damage to the endothelium is visible in OCT, to the best of our knowledge, there is no injury score available to quantify vessel damage in vivo.

The aim of this study was to establish a novel *in vivo* vascular injury score for the intraprocedural assessment of stent-induced injury as predictor of ISR using high-resolution intravascular imaging techniques like OCT. We hypothesized that severe injury caused by high-pressure stent implantation implicates a high indentation of stent struts into the vessel wall while a mild injury is associated with a lower indentation. Therefore, the stent strut indentation might reflect the actual vessel injury. Indentation was defined as the impression of the strut into the arterial wall. Furthermore, we hypothesized that higher indentation also caused increased neointimal growth. We used an established animal model for the implantation and evaluation of human coronary stents in the abdominal aorta of homozygous ApoE deficient rats as previously described (11, 12).

## Methods

### Procedural description

Seven male homozygous apolipoprotein-E-deficient (ApoE^−/-^) rats underwent stent implantation into the abdominal aorta, as described previously (11, 12). In short, stents were inserted using a trans-femoral artery access and deployed below the renal arteries in the abdominal aorta. Stents were implanted with 12 atm for 15 seconds to achieve an overstretch ratio of 1.2:1. All rats received a commercially available 2.25 × 8 mm thin-strut cobalt-chromium Multilink MiniVision stent (Abbott Vascular, St. Clara, CA).

Rats were fed a cholesterol free normal chow with 3.3% crude fat (Sniff Spezialdiäten, Soest), containing Clopidogrel (15 mg/kg; Iscover; Bristol-Myers Squibb Pharma, Uxbridge) as anticoagulant according to current European Society of Cardiology guidelines. At 4 weeks after stent implantation, rats were euthanized, and the stent-containing part of the aorta was harvested.

The animal protocol was approved by the Ministry of Nature, Environment and Country Development (Recklinghausen, Germany; AZ 87-51.04.2010.A065). The experiments were carried out in accordance with the German Animal Welfare Act (TSchG) and Directive 2010/63/EU on the protection of animals used for scientific purposes.

The human study was approved by the Ethics Committee of the Medical Faculty at RWTH (Rheinisch-Westfälischen Technischen Hochschule) Aachen (EK 062/13).

### Histological analysis

Stented aortas were harvested and fixed with 10% buffered formalin for 24 hours. Subsequently, the tissue was embedded in methylmethacrylate (Technovit 9100, Morphisto, Offenbach am Main) and processed as described previously (11, 12). In short, sequential sections of 50-70μm were cut by means of Donath’s sawing-and-grinding technique. After polishing and Giemsa staining, blinded morphological analysis was performed by an independent observer. Images of at least 3 sections each of the proximal, middle, and distal part of the stent were obtained using a Leica DMI-3000 microscope (Leica, Wetzlar, Germany), equipped with the image analysis software DISKUS (Version 4.80, C. Hilgers Technisches Büro, Königswinter, Germany). Area measurements were performed using a Tavla Pen Tablet (Braun Photo Technik, Nürnberg, Germany).

### Scoring

To obtain a new vascular indentation/injury score, we assessed the stent strut indentation for each strut in each section. Indentation (In) was defined as the impression of the strut into the vessel wall. To enable an *in vivo* assessment of indentation undisturbed by potential artifacts from struts in intracoronary imaging, indentation was not directly measured but calculated as the difference between stent strut protrusion (Pr, distance from the luminal strut edge to the point of contact with the media at the lateral sides of each strut) and strut thickness (St, 81μm in Multilink MiniVision). To confirm adequate stent expansion, the outer stent diameter was measured in all sections.

Neointimal growth after stent implantation first occurs close to the stent struts and subsequently spreads over the entire lumen as restenosis progresses. To account for this inhomogeneous distribution, neointima was measured right next to the strut (Neointima-1, NI-1) and at a distance of 40-90μm from the strut (Neointima-2, NI-2). Furthermore, we calculated neointimal thickness above the strut (NI-as) as the difference between NI-1 and Pr. The principle is visualized in a 3D reconstruction of a stented vessel in **Figure 2**.

To confirm associations between indentation and vessel wall injury, we used the Schwartz score (13) and the media compression score according to Waale (10) to assess vessel wall injury (**Figure 1**). The Schwartz score determines the degree of arterial injury by visually assessing the severity of strut penetration into the vessel wall. Scores were defined as 0 (no injury); 1 (perforation in the internal elastic membrane); 2 (perforation of the media) and 3 (perforation of the external elastic membrane to the adventitia), as described previously (13). For the Waale score, we calculated the ratio between the physiological media thickness (M-p) and the strut-compressed media thickness (M-c) for each strut in each section, as described previously (10).

**Figure 1.** **a)** Photomicrograph of stented arterial segment with histological analysis; **b and c)** schematic drawing of stented arterial segment with measuring: **b)** immediately after stenting **c)** 4 weeks after stenting; d) schematic drawing of indentation calculation. (M) media, (L) lumen, (Ni) neointima, (M-p) media-physiological, (M-c) media-compressed, (St) stent strut thickness, (Pr) protrusion, (NI-1) neointima 1 right next to the strut, (NI-2) neointima 2 at a distance of 40-90μm next to the strut, (NI-as) neointima above the strut (calculated), (In) indentation (calculated).

**Figure 2.** Visualization of the principle: μCT images and 3D reconstruction of a stented vessel with indentation of stent struts into the blue vessel wall and protrusion into the pink lumen; upper panel: cross sectional view of the vessel; lower panel: 3D visualisation of the vessel, protrusion of stent struts visible as imprints into magenta lumen reconstruction

In addition to the histopathological evaluation of stents in rats, one exemplary case of a patient was evaluated, and stent strut protrusion into the lumen was measured by OCT immediately following stent implantation.

All analyses were performed on a per strut and a per section basis. For the per section analysis, measurements of all struts from the same section were averaged.

### Statistical analysis

Statistical analysis was performed using GraphPad Prism (Version 9.0, GraphPad Software, Inc., San Diego, CA). Continuous variables are expressed as mean ± SEM. Correlations between scores and neointimal thickness as well as correlations among the scores were tested using Pearson’s R correlation coefficients. A probability value of p < 0.05 was considered statistically significant. A receiver operating characteristic (ROC) curve was used to determine the ability of stent strut indentation to predict ISR.

## Results

### Stent implantation and processing

A total of 7 stents were implanted in the abdominal aorta of ApoE^−/-^ rats. 40 sections were obtained of which 9 sections were not further analyzed due to processing failure. From a total of 323 stent struts, 42 struts without contact to the vessel wall were excluded from the analysis. In total, 31 sections and 281 struts were included in the analysis.

### Histological analysis and scoring

Per strut and per section assessments of indentation and measurements of neointima, as well as the results of the Schwartz and Waale scores, are summarized in **Table 1**. Mean Schwartz score was 0.70 ± 0.04 per strut and 0.70 ± 0.10 per section, respectively. Mean Waale score was 34.34% ± 1.24% per strut and 33.09% ± 3.44% per section, respectively. Mean stent diameter was 2.11mm ± 0.02mm. Mean indentation was 15.85μm ± 0.62μm (per strut analysis) and 14.98μm ± 1.49μm (per section analysis; **Table 1**).

**Table 1.** Average measurements:

**a) evaluation of struts**
| | Indentation<br>[$\mu\text{m}$] | NI-1 [$\mu\text{m}$] | NI-2 [$\mu\text{m}$] | NI-as<br>[$\mu\text{m}$] | Schwartz | Waale<br>[%] |
| --- | --- | --- | --- | --- | --- | --- |
| Number<br>of values | 281.00 | 281.00 | 281.00 | 281.00 | 281.00 | 281.00 |
| Minimum | 0.00 | 0.00 | 0.00 | 0.00 | 0.00 | 0.00 |
| Maximum | 81.00 | 165.00 | 140.00 | 152.00 | 3.00 | 78.60 |
| Range | 81.00 | 165.00 | 140.00 | 152.00 | 3.00 | 78.60 |
| Mean | 15.90 | 59.00 | 37.50 | 17.90 | 0.69 | 34.30 |
| Std. Error<br>of Mean | 0.62 | 2.66 | 2.05 | 1.47 | 0.04 | 1.24 |

**b) evaluation of sections**
| | Indentation<br>[$\mu\text{m}$] | NI-1 [$\mu\text{m}$] | NI-2 [$\mu\text{m}$] | NI-as<br>[$\mu\text{m}$] | Schwartz | Waale<br>[%] | stent<br>diameter<br>[mm] |
| --- | --- | --- | --- | --- | --- | --- | --- |
| Number<br>of values | 31.00 | 31.00 | 31.00 | 31.00 | 31.00 | 31.00 | 31.00 |
| Minimum | 0.10 | 0.00 | 0.00 | 0.00 | 0.00 | 0.70 | 1.94 |
| Maximum | 34.00 | 125.00 | 97.80 | 77.50 | 2.40 | 71.30 | 2.43 |
| Range | 33.90 | 125.00 | 97.80 | 77.50 | 2.40 | 70.60 | 0.49 |
| Mean | 15.00 | 55.60 | 35.50 | 17.10 | 0.70 | 33.10 | 2.11 |
| Std. Error<br>of Mean | 1.49 | 7.48 | 5.55 | 3.63 | 0.10 | 3.43 | 0.02 |

### Correlation between injury scores and neointimal thickness

The Schwartz score was positively correlated with neointimal thickness, both in the per-strut analysis (r = 0.5159 for NI-1, r = 0.4696 for NI-2 and r = 0.5207 for NI-as; all p ≤0.001) and in the per-section analysis (r = 0.7237 for NI-1, r = 0.7516 for NI-2 and r = 0.7681 for NI-as; all p ≤0.001; **Figure 3**).

**Figure 3.** Correlation matrix: **a)** evaluation of struts; **b)** evaluation of sections

Likewise, there was a positive correlation between neointimal thickness and Waale score measurements, both at strut level (r = 0.7544 for NI-1, r = 0.6216 for NI-2 and r = 0.5097 for NI-as; all p ≤0.001) and at section level (NI-1: r = 0.8737, NI-2: r = 0.7447, NI-as: r = 0.6491; all p ≤0.001; **Figures 4** and **5**).

**Figure 4.** Evaluation of stent struts: The indentation score shows a positive correlation with neointimal thickness; similar to Schwartz and Waale scores.

**Figure 5.** Evaluation of sections: Stent strut indentation was positively correlated with neointimal thickness. Similar correlations were observed between neointimal thickness and Schwartz and Waale scores.

Stent strut indentation was positively correlated with neointimal thickness, both in the per-strut analysis (r = 0.5579 for NI-1, r = 0.4087 for NI-2, r = 0.5472 for NI-as; all p ≤0.001) and in the per-section analysis (r = 0.8620 for NI-1, r = 0.7369 for NI-2 and r = 0.6871 for NI-as; all p ≤0.001; **Figures 4** and **5**).

Indentation, both at strut and at section level, showed similar accuracy for the prediction of ISR (i.e., neointimal growth) compared to the Schwartz and Waale scores (**Figure 6; Supplemental Figures 1+2**).

**Figure 6.** ROC analysis to predict neointimal thickness from indentation. AUC indicates area under curve; **a)** evaluation at strut level; **b)** evaluation at section level.

Finally, we evaluated whether *in vivo* stent strut indentation assessment was feasible in OCT in human coronary arteries. In an exemplary evaluation of post-stent OCT data of one patient, protrusion quantification was feasible, enabling the calculation of indentation *in vivo*. (**Figure 7a**).

**Figure 7.** **a**) OCT imaging of a human stented vessel with exemplary *in vivo* quantification of stent strut protrusion (left) and schematic drawing of indentation calculation (right) **b)** Photomicrograph of Giemsa-stained stented arterial segment, removed 8 years after stent implantation; adventitia (A), media (M), atherosclerotic plaque (P), calcification (C), stent struts (St), neointima (NI), lumen (L); assignment of lines: external elastic lamina (yellow), internal elastic lamina (red), atherosclerotic plaque (green), lumen (magenta); lower left part of the section: high injury and indentation, internal elastic lamina lacerated (**), upper right part of the section: lower injury and indentation, no laceration of the internal elastic lamina (*).

In an exemplary histopathological evaluation of a human stented coronary artery, sections with pronounced indentation also showed profound signs of vessel wall injury, as shown in **Figure 7b**.

## Discussion

In this study, we suggest a novel *in vivo* injury score to predict restenosis risk after coronary stent implantation. As validated by *postmortem* histopathology in a rat model of stent implantation, the score highly consistently correlates with neointimal thickness by taking into account to which degree the stent struts are embedded or indented into the vessel wall at time of implantation. This study demonstrates the indentation score, a new *in-vivo* vascular injury score as a suitable tool for *in-vivo* assessment and quantification of the stent induced injury directly after stent implantation and for *in-vivo* prediction of restenosis development thus enabling stent implantation optimization.

Iatrogenic vessel wall injury during PCI remains inevitable and is a challenge hard to control for the physician. Stent strut indentation can be influenced by the stent size, implantation pressure or through re-dilatation or proximal optimization technique. The characteristics and properties of the plaque as well as its inhomogeneous distribution along the vessel wall, the amount of calcification and consequently plaque rigidity are important factors to be taken into consideration before PCI.

A pronounced injury and consequently an increased indentation will lead to a stronger overstretch of the wall vessel with formation of a thicker neointima and stenosis. Implantation of a smaller stent or stent dilatation with lower pressure to avoid excessive media injury with concomitant restoration of proper blood flow can lead to malapposition and thrombosis with fatal consequences.

Here, for validation of a novel *in-vivo* score, we used a rat model of stent-induced vessel injury. As neointima development after vessel injury is accelerated in rats and is known to be near-complete after 4 weeks, we used a 4-week time point for planimetry. We demonstrated that the correlation of the novel stent strut indentation score with vessel injury is comparable to established histological scores and highly positively correlates with neointimal thickness development.

OCT is an intravital high resolution imaging modality that provides detailed visualization of vessel wall injury during stent implantation *in vivo*, thus enabling an intravital assessment of the novel indentation score. In general, current practice of OCT-derived stent optimization is based on MSA cutoff or malapposition control, while a distinct post-implantation score to characterize the procedure’s results at the individual patient’s level is missing. Thus, the assessment of the novel indentation score might be useful for individual lesion treatment and evaluation of stent apposition and could therefore potentially provide a future tool for patient-individualized PCI.

## Limitations

Stent implantation was performed in the abdominal aorta of ApoE knockout rats. In contrast to human coronary arteries which are muscular vessels, the abdominal aorta is of the elastic type and therefore differs in its structure and response to injury. Furthermore, the stents were not implanted into a stenotic vessel as it would be the case in human stent implantation. However, the ApoE^−/-^ rat model has proven to be a suitable model to approximate the conditions in the human atherogenic vessel. Also, indentation was quantified after formalin fixation of the vessel, thus, formalin-related tissue shrinkage cannot be ruled out.

## Conclusions

Stent strut indentation was positively correlated with vessel wall injury and subsequently with neointimal growth in histology. As it is feasible to assess stent strut indentation *in vivo* by means of OCT imaging, our findings have a potentially important clinical impact. In conventional stent implantation with targeting a maximization of MSA, injury to the deep vessel wall as a result of aggressive stent expansion leads to greater ISR than a moderate stent expansion. An *in-vivo* injury score enables the periprocedural assessment of stent-induced damage and therefore allows for an optimization of the stent implantation process. After initial moderate stent expansion, the MSA can be carefully increased by post dilation, and a balance between maximizing the final MSA and minimizing mechanical damage in the vessel wall can be achieved. Therefore, the assessment of stent strut indentation has the potential to become a valuable tool in daily practice.

## Data Availability

All data produced in the present study are available upon reasonable request to the authors

## Acknowledgements

Marek Weiler (Institute for Experimental Molecular Imaging, Medical Faculty, Rheinisch-Westfälische Technische Hochschule (RWTH) Aachen University Clinic, 52074, Aachen, Germany) performed the μCT Scan.

## Funding

This work was supported by the German Research Foundation (DFG) [grant numbers 465213526; 395712048].

## Disclosures

None.

## Notes

### Competing Interest Statement

The authors have declared no competing interest.

### Author Declarations

Ethics committee of Medical Faculty at Rheinisch-Westfaelische Technischen Hochschule Aachen gave ethical approval for this work (EK 062/13)

