## Supplemental Figures 1 and 2 for "Development and validation of a novel *in-vivo* vascular injury score for prediction of in-stent restenosis"

**Corresponding Author:**

Dr. med. Anne Cornelissen

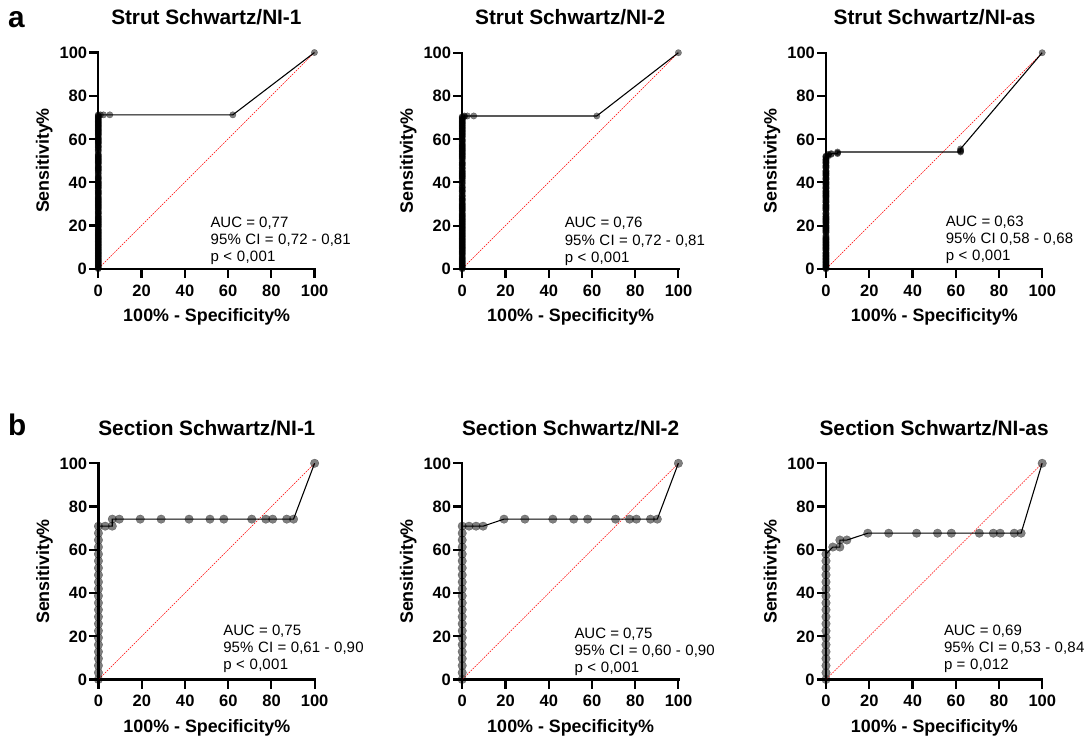

**Supplemental Figure 1.** ROC analysis to predict neointimal thickness from Schwartz score. AUC indicates area under curve; **a)** evaluation at strut level; **b)** evaluation at section level.

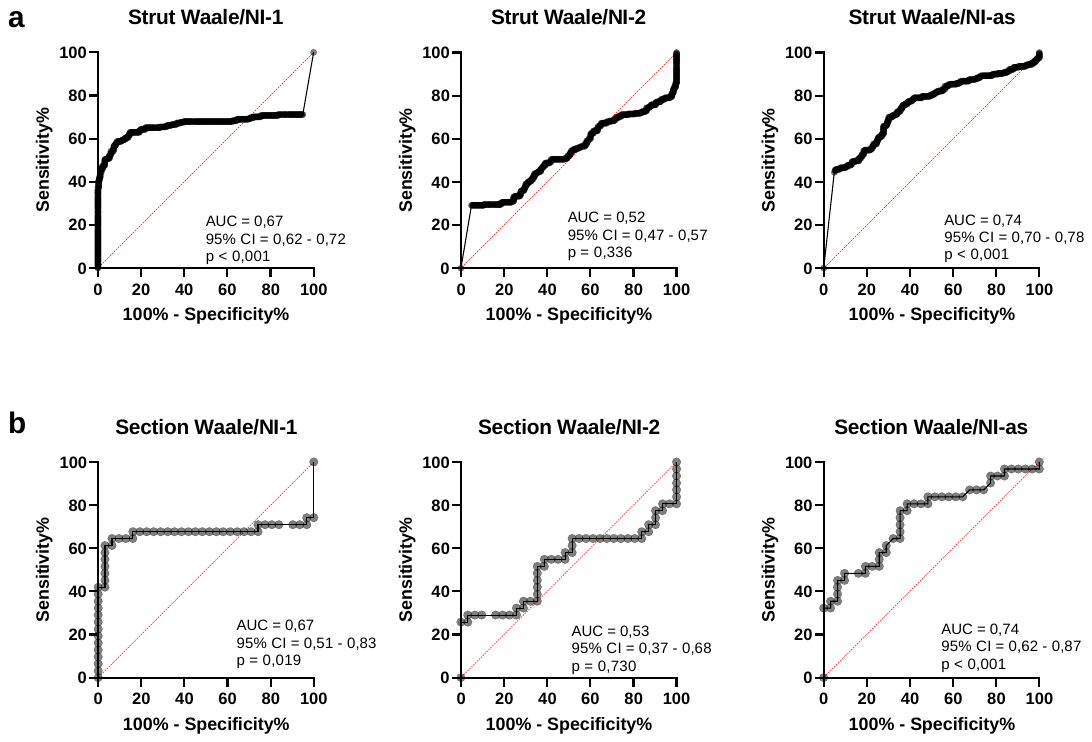

**Supplemental Figure 2.** ROC analysis to predict neointimal thickness from Waale score. AUC indicates area under curve; **a)** evaluation at strut level; **b)** evaluation at section level.
